# Estimating the burden of mpox among MSM in South Africa

**DOI:** 10.1101/2024.08.13.24311919

**Authors:** Ruth McCabe, Leigh F. Johnson, Lilith Whittles

## Abstract

Despite seeing few cases during the 2022-23 mpox global outbreak, recent reports of 22 cases among men-who-have-sex-with-men (MSM) in South Africa, including three deaths, have raised concerns about underreported community transmission. We used a Monte Carlo simulation model to estimate the true epidemic size, considering increased severity of mpox among MSM living with advanced HIV (MSMLAHIV), documented overrepresentation of people living with HIV (PLHIV) among mpox cases, and HIV prevalence in South Africa. We estimate there have been between 220-450 cases among MSM LHIV in South Africa, implying a total of 290-560 cases among all MSM. We provide an upper bound of 750-1,600 cases as a sensitivity analysis where the prevalence of HIV among mpox patients is the same as population prevalence among MSM in South Africa. Estimates in both scenarios suggest a substantial number of undetected cases, with case ascertainment rates estimated between 1% and 8%. Our findings underscore the need for enhanced surveillance, targeted public health interventions, and awareness campaigns to mitigate the outbreak’s impact at a population-level.

## Introduction

Since 2022, the global outbreak of mpox has caused over 97,000 cases across 116 countries (1). In contrast to outbreaks in historically endemic countries, transmission mostly occurred via sexual networks, with cases heavily concentrated among men who have sex with men (MSM). Prior to 2024, South Africa was largely unaffected by the global outbreak of mpox, recording only five cases, all of which were linked to international travel (2). However, between May and July 2024 the country has reported 22 cases and three deaths (2). All 22 cases occurred in men aged between 17 and 43 (3), most of whom self-identify as MSM (4). Genomic sequencing was performed on a subset of five cases, all of which were confirmed to be MPXV subclade IIb – linking them to the 2022-23 global outbreak, rather than the ongoing epidemic of MPXV clade I in the Democratic Republic of Congo and bordering countries (5). The global outbreak of MPXV clade IIb has been associated with a low case fatality ratio (CFR) 0.21% (n=203/97,745; 95% CI: 0.15% - 0.24%), so the three recent deaths in South Africa have prompted concern about the true burden of mpox. Furthermore, the recent cases had no history of international travel or attendance at high-risk gatherings, and are largely without epidemiological links, fuelling concern of sustained, undetected community transmission (4,6).

Globally, PLHIV have been disproportionately affected by mpox, and are over-represented among both cases and deaths (7). Increased mpox severity and fatality rates among people living with advanced HIV (PLAHIV) have been well-documented (8), which is of particular concern due to typically higher prevalence of HIV among MSM compared to the general population. South Africa is among the countries most affected by the HIV worldwide, with an estimated 18.3% (95% CI: 15.6%-20.5%) of the adult population living with HIV, which rises to 29.4% (25.5% - 33.5%) in MSM (9,10). At least two-thirds of the recent South African mpox cases (n=15/22) occurred in people living with advanced HIV – including the individuals with fatal outcomes (6).

Here, by leveraging the relationship between deaths and cases via the CFR, we estimate the potential total number of mpox cases underlying the observed number of cases and deaths thus far (July 2024), accounting for the high prevalence of HIV in South Africa and its effect of mpox severity.

## Methods

To estimate the total number of mpox cases in MSM, *C*, given observed deaths, all of which were in MSMLAHIV (*D* = *D*_*AHIV*_), we first calculated the expected number of cases among MSMLAHIV, *C*_*AHIV*_, using the corresponding observed mpox CFR among PLAHIV, *P* (*D*_*AHIV*_ |*C*_*AHIV*_):

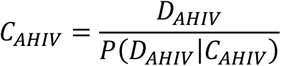

We then used this estimate to calculate the expected number of cases among all MSMLHIV, *C*_*HIV*_, by considering the proportion of mpox cases with HIV whose infection is in an advanced stage:

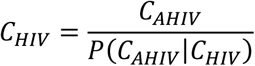

Finally, we estimated the total number of mpox cases among MSM, using the prevalence of HIV among mpox cases:

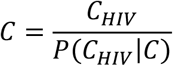

### Model parametrisation

#### Case fatality ratio in MSMLAHIV

A global case series of mpox among PLHIV found clear evidence of higher CFRs in people with lower CD4+ T-cell counts (8). Of the 27 mpox deaths recorded among a sample of 382 cases with HIV coinfection during the 2022-23 global outbreak, 23 occurred in the 85 individuals CD4+ T-cell count <100/mm^3^, corresponding to a CFR of 27.1% (95% exact binomial confidence interval (CI): 18.0% -37.8%). The remaining four deaths were recorded in the 94 individuals with CD4+ T-cell counts between 100-200/mm^3^, equating to a CFR of 15.1% (95% CI: 10.2% - 21.2%) in those with CD4+ T-cell count <200/mm^3^. No deaths occurred in the 204/382 mpox cases with CD4+ T-cell count ≥200/mm^3^. The definition of ‘advanced HIV’ used in the South African mpox case reports is unclear but is typically defined as CD4+ T-cell count <200/mm^3^, which we take as our central value. However, we perform sensitivity analysis using an alternative definition based on CD4+ T-cell count <100/mm^3^.

#### Proportion of mpox cases LHIV with advanced HIV

We assume that the proportion of MSMLHIV whose HIV infection is at an advanced stage is the same among mpox cases as in the MSM population, specifically, that the probability of having advanced HIV can be considered independent of the probability of having mpox, conditional on HIV infection. To quantify the proportion of MSM LHIV whose infection is in an advanced stage, we use estimates from the Thembisa 4.7, a transmission-dynamic deterministic compartmental model of HIV in South Africa (9), which has been used to produce HIV estimates for national policy development including those reported by UNAIDS (11). The model predicts that 6.6% of adults living with HIV in South Africa in 2024 will have a CD4+ T-cell count below 200/mm^3^.

For our sensitivity analysis of the definition of ‘advanced HIV’, we assume that 3.3% of the population will have CD4+ T-cell count <100/mm^3^, (i.e. that CD4+ T-cell counts are distributed roughly uniformly below 200/mm^3^). This assumption aligns with the global case series of mpox in PLAHIV, in which mpox cases with CD4+ T-cell counts <100/mm^3^ comprised 47.4% (n=84/179; 95% CI 40.0% - 55.1%) of mpox cases with CD4+ T-cell counts<200/mm^3^ (8).

#### Proportion of mpox cases LHIV

Globally, PLHIV have observed a higher incidence of mpox cases, likely due to shared sexual network risk (7). A conservative lower bound for the prevalence of HIV among mpox cases would therefore be the prevalence of HIV among MSM, *P* (*N*_*HIV*_), which the Thembisa model estimates at 29.5% (95% CI: 25.5% - 33.5%) in South Africa. However, this method may overstate the overall number of cases and so we consider this as a sensitivity analysis. Instead, our central scenario considers that the odds of LHIV given mpox infection are proportional to the odds of LHIV.

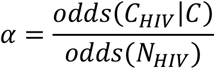

We estimated the odds ratio, *α*, using mpox case data from the USA in which HIV prevalence was 56.4% (n=5,350/9,492; 95% CI: 55.4% - 57.4%) (12), compared to prevalence of 12.4% (95% CI: 10.9% - 13.8%) among all MSM (13). Serological evidence has shown a low prevalence of undiagnosed mpox in the US, which supports the use of the data to form an unbiased assessment of risk-patterns (14). These data suggest the odds of having HIV were *α* = 9.2 (95% CI: 7.9 – 10.6) times higher among mpox cases than in the general MSM population.

We multiplied the odds of HIV infection among all MSM by *α* to obtain a central estimate of the prevalence of HIV among mpox cases in South Africa:

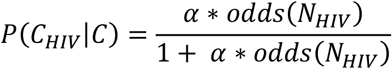

#### Modelled scenarios

We created a Monte Carlo simulation model to quantify the potential number of mpox cases in South Africa i) in MSMLAHIV; ii) in all MSMLHIV; and iii) in MSM in total. Conditional probabilities were modelled using beta distributions, parameterised from the literature, either directly from reported sample numbers where possible, or estimated via method-of-moments from mean and 95% confidence intervals (Table 1). We generated 10,000 samples from parametric distributions to propagate the statistical uncertainty inherent in the parameter estimates throughout the calculations, allowing us to present results in terms of mean and 95% confidence intervals. Analyses were performed in R v4.2.2; model code is available at https://github.com/mrc-ide/mpox_sa.

**Table 1:** Model parameters, descriptions and sources.

| Parameter | Description | Distribution | Mean (95% CI) | Source |
| --- | --- | --- | --- | --- |
| $P(D\vert C_{CD4<200})$ | Mpox CFR among PLAHIV | B(27, 152) | 17.8%<br>(10.2% - 20.7%) | (8) |
| $P(HIV_{SA})$ | HIV prevalence among MSM in South Africa | B(146, 351) | 29.4%<br>(25.5% - 33.5%) | Thembisa v4.7(9);<br>Beta distribution fitted<br>via MOM |
| $P(HIV_{US})$ | HIV prevalence among MSM in USA | B(241, 1706) | 12.4%<br>(10.9% - 13.9%) | (13); Beta distribution<br>fitted via MOM |
| $P(HIV_{US}\vert C)$ | HIV prevalence among MSM with mpox in USA | B(5350, 4142) | 56.4%<br>(55.3% - 57.3%) | (12) |
| $\alpha$ | Odds ratio of LHIV among mpox cases vs. all MSM | NA | 9.2<br>(8.0 – 10.6) | Estimated from USA case data |
| **Sensitivity analysis** |  |  |  |  |
| $P(D\vert C_{CD4<100})$ | Mpox CFR among PLAHIV | B(23, 62) | 37.1%<br>(18.2% - 36.9%) | (8) |
| $\alpha$ | Odds ratio of LHIV among mpox cases vs. all MSM | NA | 1 | NA |

## Results

Table 2 summarises the results of the central scenario and associated sensitivity analyses.

**Table 2:** Estimated potential number of mpox cases in South Africa among MSMLAHIV, MSMLHIV, and in total under four scenarios with corresponding parameter assumptions. Results are presented as mean (95% confidence interval) where applicable

| Scenario: | Central | Sensitivity Analyses |  |  |
| --- | --- | --- | --- | --- |
| Odds ratio of LHIV among mpox cases vs. all MSM: | 9.2 (8.0 – 10.6) | 9.2 (8.0 – 10.6) | 1 | 1 |
| Definition of AHIV: | CD4 <200/mm<sup>3</sup> | CD4 <100/mm<sup>3</sup> | CD4 <200/mm<sup>3</sup> | CD4 <100/mm<sup>3</sup> |
| Observed $D_{AHIV}$ | 3 | 3 | 3 | 3 |
| Assumed $P(D_{AHIV}\vert C_{AHIV})$ | 15.0%<br>(10.2% - 20.6%) | 27.1%<br>(18.1% - 36.8%) | 15.0%<br>(10.2% - 20.6%) | 27.1%<br>(18.1% - 36.8%) |
| Estimated $C_{AHIV}$ | 21 (15 - 29) | 11 (8 - 17) | 21 (15 - 29) | 11 (8 - 17) |
| Observed $C_{AHIV}$ | 15 | 15 | 15 | 15 |
| Assumed $P(C_{AHIV}\vert C_{HIV})$ | 6.6% | 3.3% | 6.6% | 3.3% |
| Estimated $C_{HIV}$ | 313<br>(227 - 444) | 458<br>(455 - 503) | 313<br>(227 - 444) | 458<br>(455 - 503) |
| Assumed $P(C_{HIV}\vert C)$ | 79.1%<br>(75.0% - 82.9%) | 79.1%<br>(75.0% - 82.9%) | 29.4%<br>(25.5% - 33.5%) | 29.4%<br>(25.5% - 33.5%) |
| Estimated $C$ | 395<br>(287 - 562) | 579<br>(549 - 640) | 1,068<br>(750 - 1,543) | 1,565<br>(1,361 - 1,832) |
| Estimated $P(D\vert C)$ | 0.78%<br>(0.53% - 1.05%) | 0.52%<br>(0.47% - 0.55%) | 0.29%<br>(0.19% - 0.40%) | 0.19%<br>(0.16% - 0.22%) |
| Estimated cases detected | 5.7%<br>(3.9% - 7.7%) | 3.8%<br>(3.4% - 4.0%) | 2.1%<br>(1.4% - 2.9%) | 1.4%<br>(1.2% - 1.6%) |

### Estimated mpox cases among MSM LHIV

The three mpox deaths observed among PLAHIV in South Africa are consistent with an underlying burden of 21 cases (95% CI: 15 – 29) (Table 2). The 15 cases that have been observed in PLAHIV up to June 2024 are therefore broadly consistent with modelled expectations, subject to case under-ascertainment. Taking the observed cases as a lower bound of the true total implies a case-ascertainment rate of 74.9% (95% CI: 51.2% - 100%) among MSMLAHIV. Under the assumption that 6.6% of HIV-positive MSM with mpox have CD4+ T-cell counts < 200/mm^3^, we estimate total cases in PLHIV to be 313 (95% CI: 227 – 444).

### Estimated total mpox cases in MSM

Allowing for overrepresentation of PLHIV among mpox cases, in line with relative odds observed in the US, suggests an HIV prevalence of 79.1% (95% CI: 74.9% - 82.9%) among MSM with mpox in South Africa, compared to 29.4% (95% CI: 25.5% - 33.5%) in all MSM. Taking this population prevalence of HIV into account, we estimate the total burden of mpox at 395 (95% CI: 287 - 562) cases. These estimates correspond to CFR at the high end of that observed in the global outbreak: 0.5%-1.0%, reflecting the high burden of HIV in South Africa, and an mpox case ascertainment of around 3.9%-7.7% to date.

### Sensitivity analyses

Using the alternative definition of ‘advanced HIV’ as CD4+ T-cell count <100/mm^3^, the observed three deaths are compatible with 11 cases (95% CI: 8 - 17) among MSMLAHIV, which would imply that case ascertainment among MSMLAHIV has been extremely high, with 99.4% (95% CI: 90.4% - 100%) of infections in this group already observed. Taking the 15 observed cases in PLAHIV as a lower bound for the true number of cases, the assumption that 3.3% of MSMLHIV have CD4+ T-cell counts <100/mm^3^ is compatible with a higher number of cases than the CD4+ T-cell count <200/mm^3^ definition, suggesting an underlying case burden of 458 (95% CI: 455 – 503) among MSMLHIV and 579 (95% CI 549 - 640) in total.

In the conservative scenario, where we assume that prevalence of HIV among mpox cases is in line with background HIV prevalence among MSM in South Africa at 29.4% (95% CI: 25.5% - 33.5%), we estimate the total number of cases in the outbreak to be 1,068 (95% CI: 750 - 1,543) if the reported deaths had CD4+ T-cell counts <200/mm^3^, or up to 1,565 (95% CI: 1,361 - 1,832) under the stricter definition of CD4+ T-cell counts <100/mm^3^. These figures are 2-3-fold higher than in the central scenario and are consistent with an overall infection fatality ratio of around 0.2-0.3%, similar to that observed globally, and suggest that only 1%-3% of recent cases in South Africa have been detected.

## Discussion

Using recent reports of 15 mpox cases and three deaths among MSMLAHIV in South Africa, we used a simple probabilistic model to estimate the underlying burden of unobserved cases, both in total and among MSMLHIV.

A key advantage of our pragmatic approach is that we extrapolate from published literature on mpox epidemics in similar populations, as well as incorporating context-specific factors, such as the high prevalence of HIV among MSM in South Africa. A further strength is that we incorporate parametric uncertainty via the use of Monte Carlo simulations, allowing us to present a comprehensive view of the range of outcomes compatible with the observed number of cases and deaths. Finally, we present plausible upper and lower bounds for the true burden of mpox in South Africa by conducting a range of sensitivity analyses; we use different definitions of advanced HIV based on CD4 count thresholds, and present both cautious and neutral assumptions about the prevalence of HIV among mpox cases relative to the general population.

To enable estimation of the mpox case burden, we have made simplifying assumptions that should be borne in mind when interpreting the results. The uncertainty associated with each of these assumptions has been propagated through to our final estimates, which explains the relatively large confidence intervals that we report here. Firstly, the accuracy of our results is dependent on any biases in published parameter estimates; for example, if many cases among PLAHIV have been undetected, then the CFR could be lower than estimated in the global case series (8), which would serve to increase estimates of case numbers presented here. Concerns around biased estimates of severity in PLAHIV are mitigated by the fact that mpox symptoms are also more severe in this population, suggesting ascertainment is likely high (8). However, it could also be the case that the true number of deaths from mpox is under-reported due to ambiguity in the documentation of causes of death, which may suggest the observed CFR is an under-estimate. Secondly, given the case patterns observed globally, as well as recently in South Africa, our study limits the population at risk to MSM, and does not consider transmission to other populations, such as female partners. Consequently, our estimates may be an underestimate of the full burden of mpox in South Africa. Thirdly, we assume the severity of clade IIb mpox is unchanged since global outbreak, and that national differences in the observed CFR are attributable to the underlying prevalence of AHIV among MSM. Fourth, we assume that prevalence of AHIV is the same in mpox patients as in among all MSM LWHIV – however individuals with advanced HIV may be less likely to engage in sex, and so be less likely to contract mpox, compared to other HIV-infected individuals, which would also serve to increase estimates of the underlying number of cases. Moreover, recent studies have suggested that the percentage of men LAHIV in Sub-Saharan Africa could be as high as 13% (15). Adopting this higher estimate in our model would approximately halve the expected number of mpox cases. Conversely, LAHIV may make contracting mpox more likely if sexual contact with an infected individual does occur, due to inhibited mucosal immunity. Finally, we note that while information on the HIV status on the initial 20 cases was reported, this was not mentioned for the most recent two cases. We assume that these individuals are not LAHIV but note that if this were true then the estimated number of mpox cases among MSM would increase.

To place our findings in the broader context of global mpox outbreaks among MSM, in countries where epidemics have risen and peaked, likely due to a combination of network saturation effects and behavioural adaptation, the final epidemic size has been estimated to range from 0.1% to 1.5% of the MSM population (16), noting that determining accurate population denominators for MSM can be challenging. In South Africa, the MSM population is estimated at approximately 310,000 individuals (10), suggesting the potential final size of the mpox outbreak could reach up to 4,650 cases, or 3-10x the current estimated size. This underscores the urgency for targeted public health interventions, surveillance and risk communication and communication engagement (RCCE) to manage the outbreak effectively and mitigate its impact.

## Conclusions

A combination of evidence, including 22 mpox cases with no reported travel history, no epidemiological links, and no attendance at high-risk events/venues, coupled with three observed deaths, despite the low case fatality reported globally, suggests undetected community transmission of mpox in South African MSM. We estimate a current epidemic size of between 280-570 cases in our central scenario, or 750-1,600 cases in the conservative scenario assuming PLHIV are not overrepresented among mpox cases – corresponding to a detection rate of between 1 in 14 and 1 in 100 cases. The true size of the epidemic could be confirmed with serological studies, but in the meantime, awareness campaigns to encourage testing, symptomatic abstention from sex, and vaccination where available are recommended to mitigate the risks of future transmission.

## Data Availability

Analyses were performed in R v4.2.2; model code is available at https://github.com/mrc-ide/mpox_sa. All data are publicly available and cited in the manuscript.

## Contributions

LKW conceived the study, designed the methodology, and performed the analysis. LKW and RM drafted the report. LFJ advised on parameter values and revised the report.

## License

For the purpose of open access, the author has applied a ‘Creative Commons Attribution (CC BY) licence to any Author Accepted Manuscript version arising.

## Acknowledgments

LKW would like to acknowledge Dr Alyssa Bilinski’s ‘Napkin Math’ approach, which greatly guided her thinking.

